# Anosmia and other SARS-CoV-2 positive test-associated symptoms, across three national, digital surveillance platforms as the COVID-19 pandemic and response unfolded: an observation study

**DOI:** 10.1101/2020.12.15.20248096

**Authors:** Carole H. Sudre, Ayya Keshet, Mark S. Graham, Amit D. Joshi, Smadar Shilo, Hagai Rossman, Benjamin Murray, Erika Molteni, Kerstin Klaser, Liane D Canas, Michela Antonelli, Marc Modat, Joan Capdevila Pujol, Sajaysurya Ganesh, Jonathan Wolf, Tomer Meir, Andrew T. Chan, Claire J. Steves, Tim D. Spector, John S. Brownstein, Eran Segal, Sebastien Ourselin, Christina M. Astley

## Abstract

**Background:** Multiple participatory surveillance platforms were developed across the world in response to the COVID-19 pandemic, providing a real-time understanding of community-wide COVID-19 epidemiology. During this time, testing criteria broadened and healthcare policies matured. We sought to test whether there were consistent associations of symptoms with SARS-CoV-2 test status across three national surveillance platforms, during periods of testing and policy changes, and whether inconsistencies could better inform our understanding and future studies as the COVID-19 pandemic progresses.

**Methods:** Four months (1st April 2020 to 31st July 2020) of observation through three volunteer COVID-19 digital surveillance platforms targeting communities in three countries (Israel, United Kingdom, and United States). Logistic regression of self-reported symptom on self-reported SARS-CoV-2 test status (or test access), adjusted for age and sex, in each of the study cohorts. Odds ratios over time were compared to known changes in testing policies and fluctuations in COVID-19 incidence.

**Findings:** Anosmia/ageusia was the strongest, most consistent symptom associated with a positive COVID-19 test, based on 658,325 tests (5% positive) from over 10 million respondents in three digital surveillance platforms using longitudinal and cross-sectional survey methodologies. During higher-incidence periods with broader testing criteria, core COVID-19 symptoms were more strongly associated with test status. Lower incidence periods had, overall, larger confidence intervals.

**Interpretation:** The strong association of anosmia/ageusia with self-reported SARS-CoV-2 test positivity is omnipresent, supporting its validity as a reliable COVID-19 signal, regardless of the participatory surveillance platform or testing policy. This analysis highlights that precise effect estimates, as well as an understanding of test access patterns to interpret differences, are best done only when incidence is high. These findings strongly support the need for testing access to be as open as possible both for real-time epidemiologic investigation and public health utility.

**Funding:** NIH, NIHR, Alzheimer’s Society, Wellcome Trust

**Research in context:** *Evidence before this study:* As the COVID-19 pandemic has evolved, testing capacity expanded and governmental guidelines adapted, generally encouraging testing with a broader set of symptoms, not just fever with respiratory symptoms. In parallel, multiple large-scale citizen science digital surveillance platforms launched to complement knowledge from laboratory and somewhat smaller clinical studies. Symptoms such as loss of sense of smell have been identified as strongly predictive of COVID-19 infection in both clinical and syndromic surveillance analyses, and have therefore been used to inform these testing policy changes and access expansion.

*Added value of this study:* This study identifies symptoms that are or are not consistently associated with SARS-CoV-2 test positivity over time and across three country-based COVID-19 surveillance platforms in the United States, United Kingdom and Israel. These platforms are website and smartphone based, as well as cross-sectional and longitudinal. The study period of 4 months covers fluctuating COVID-19 prevalence during the fall of the first wave and, in some areas, rise of the second wave. In addition, the study period overlaps expansion of test access and test seeking. Importantly, these analyses track and highlight the value of individual symptoms to predict SARS-CoV-2 test positivity under a range of conditions.

*Implications of all the available evidence:* Despite differences in surveillance methodology, access to SARS-CoV-2 testing and disease prevalence, loss of sense of smell or taste was consistently the strongest predictor of COVID-19 infection across all platforms over time. As access to testing broadened, the relevance of COVID-like symptoms and consistency of their predictive ability became apparent. However, confidence bounds generally widened with a fall in COVID-19 incidence. Therefore, for the most robust symptom-based COVID-19 prediction models should consider surveillance data during periods of higher incidence and improved test access, and effect estimates that replicate across different epidemiologic conditions and platforms.

## Introduction

Participatory syndromic surveillance has informed public health for nearly a decade^1,2^, though it was the COVID-19 pandemic that spurred the rapid development of multiple, digital monitoring platforms^3–9^ to accelerate our understanding of and response to SARS-CoV-2 globally^10^. These citizen science initiatives encompass a range of participant interfaces including website^3,5,9^, phone^5^, text message^9^ and smartphone applications^4,6^, using cross-sectional and longitudinal study designs, and implementing varying degrees of population sampling or engagement.

Real-time, community-based data from these platforms are strongly complementary to the so-called “hard outcomes” — i.e. COVID-19 cases, hospitalizations and deaths^11^ — particularly in the setting of inadequate testing, delayed or absent reporting, or when ascertained outcomes are only among the most severe cases (e.g. clinical features of hospitalized COVID-19)^12,13^. As an example of the utility of such platforms, prediction of COVID-19 infection using symptom-based scores was pioneered using data from these platforms in response to the limited testing capacity early during the pandemic, and highlighted early the potential importance of smell and taste disorders^8,14^.

COVID-19 participatory surveillance platforms function in regions that have been variably impacted by the pandemic, though there has been no direct comparison of these data. Testing policies^15^, test access^16^, and Covid-Like-Illness (CLI) definitions have also varied substantially from country-to-country and over time. In many regions, testing was primarily targeted at those whose symptoms (or exposures) met strict criteria (e.g. fever and respiratory symptoms)^17^, and then later CLI symptoms were broadened to acknowledge the spectrum of COVID-19 presentations^18^, and to include other, sometimes highly specific features (e.g. anosmia)^19^.

With all of these spatio-temporal changes in policies and access, as well as platform-specific study design features, we sought to identify which symptoms were consistently associated with SARS-CoV-2 test positivity — and thus might represent the most clinically and epidemiologically relevant COVID-19 signals. In addition, we hypothesized that the predictive power of other symptoms evolved over time, in concert with country-specific testing practices and disease prevalence. Towards this end, we undertook a comparison of the association of putative CLI symptoms with self-reported SARS-CoV-2 testing results, over time, in three countries with different testing policies, and across three citizen-science digital surveillance platforms.

## Methods

Briefly, data from three country-based participatory surveillance platforms, spanning a four-month period of observation early in the pandemic (1st April 2020 to 31st July 2020), were used to estimate odds ratios (OR) for symptoms on self-reported SARS-CoV-2 test positivity among non-healthcare workers (as healthcare workers generally received different access to testing). Specifics of each cohort from Israel, the United Kingdom (UK), and the United States (US), as well as exposures, outcomes, and statistical analysis are in prior publications and summarized hereafter. Mapping of survey questions across platforms in Supplementary table 1.

## Study Populations

### Facebook/Carnegie Mellon University US COVID-19 Symptom Survey (US)

This research is based on survey results from Carnegie Mellon University’s Delphi Research Group. The US Facebook COVID-19 Symptom Survey hosted by the Carnegie Mellon Delphi Research Center provided web-based surveys to Facebook users^20^. Surveys asked about geographic location, age, sex, working in a healthcare setting in the prior 5 days before survey, and the presence of symptoms in the prior 24 hours. Symptomatic respondents were additionally asked about SARS-CoV-2 test seeking, test access and test results. From launch 6th April 2020 through 31st July 2020, there were N=6,626,897 anonymous surveys, presumed to be from unique respondents, from 50 US states and the District of Columbia. Ssurvey sampling strategies are used to increase representativeness of the US population by sampling from the Facebook active user base and raking across US census age, sex and geographic region to develop survey weights. Please refer to data documentation regarding survey sampling methods^20^. Primary analyses across all cohorts represent unweighted parameters, as survey weights were not available in the UK and Israel. Survey-weighted estimates are nearly identical. Supplementary figure 2 and 4 include sensitivity analyses with survey weights, with filtering by symptom response patterns (as a proxy for survey misuse, recall bias or severity) and with subsetting on self-reported illness duration. This study was approved by the Boston Children’s Hospital IRB (P00023700).

### UK Zoe Covid Symptom Study App (UK)

The data used for this work was collected through the COVID Symptom Study App, developed by Zoe Global Limited with input from physicians and scientists from King’s College London, Massachusetts General Hospital, Lund and Uppsala Universities^4^. The app was launched in the UK on March 24th 2020 and at 31st July counted 3,360,949 adult participants in the UK. At registration, users are asked for personal characteristics (age, gender, and whether they are a healthcare worker). App users are asked to report their health status everyday indicating their symptoms, if they experience any. In addition, they record their test results for COVID-19. Research studies on data collected through the app are approved by King’s College London Ethics Committee REMAS ID 18210, review reference LRS-19/20-18210 and all participants provided consent. Thanks to a partnership between the Department for Health and Social Care, tests were made available to users of the app upon invitation from the app maintainers (Zoe) from the 26 April 2020. See Supplementary figure 1 and 3 for a sensitivity analysis of outcomes stratified by test invitation (to assess the role of test seeking behavior) or using symptom windows that extend beyond the test date (to compare to the cross-sectional US data). All tests results were analyzed in the main analysis. Multiple tests per user were censored within the symptom window following the test, or once a test resulted positive.

### Corona Israel (Israel)

The Israel Corona study was collected through an online survey (https://coronaisrael.org/) that included a one-minute, anonymous, online questionnaire. From the date first published ^5^ on 14th March 2020 through 31st July 2020, there were N=131,799 completed surveys (15,550 unique users). Survey responses were collected directly through the online platform. Responders were asked to report information on age, gender, geographic location, prior medical conditions and whether they are a healthcare worker as well as symptoms experienced in the prior 24-hours for themselves and for each member of the family. Additionally, SARS-CoV-2 testing and test results were reported. This study was approved by the Weizmann Institute of Science review board (IRB). The IRB waived informed consent as all identifying information was removed prior to analysis.

### Study period, inclusion and exclusion criteria

Data from 1st April 2020 (or first testing data acquisition, if later) through 31st July 2020 were aggregated into weeks starting Mondays. The study population was restricted to respondents self-reporting baseline age 18 to 100 years (US age category > 75 years inclusive), sex male or female and non-healthcare workers. US survey age-bins assigned age was the included decade (e.g. 12-24 as 20 years, 35-44 as 40 years, >75 as 80 years). Users with missing demographic data were excluded.

### Exposures (symptoms) and outcomes (COVID-19 test status)

Eleven symptoms shared across at least two platforms were grouped into meta-symptoms (e.g. myalgias/arthralgias inclusive of muscle pain, joint pain in Supplementary table 1). Symptoms that were shared but had limited responses, and thus could not be compared (i.e. abdominal pain, rash, confusion), were excluded. Self-reported symptoms were considered present if logged within 14 days prior to the COVID-19 test (Israel, UK), or if logged as present in the prior 24 hours (US), which is also when the test was reported. Sensitivity analyses of UK data to show the impact of including symptoms following testing (as in the US), and of US data to show the impact of illness duration are shown in Supplementary figure 3 and 4, respectively.

The primary outcome was the self-reported result of the SARS-CoV-2 test (i.e. positive versus negative). A secondary outcome (UK, Israel) included whether testing was completed (i.e. test obtained versus not). In the US platform, secondary outcomes were whether testing was sought (i.e. attempted to test versus did not), and whether testing was obtained (i.e. among those attempting testing, able to test versus not). Testing counts and positive test proportions were tabulated as the number of users (UK) or surveys (US, Israel) and the ratio of test positives to total resulted tests. multiple test results could be reported (Israel, UK), if tests were performed at less than 14 day window, only the first test was considered. Users were censored for a 14 day window, or after a first positive test.

### Statistical analysis

Logistic regression of each symptom (binary) on SARS-CoV-2 test status (binary) adjusted for age (continuous) and sex (binary) was performed separately in each cohorts, for each moving 5-week window, plotted on the last week of the interval. Association tests for secondary outcomes were conducted in a similar manner. Israel’s primary analyses aggregated across the entire follow-up period due to the smaller sample size. Supplementary figure 5 is a sensitivity analysis of 5-, 7- and 9-week aggregation windows for Israel. Analyses were performed using (R 3.6.3 glm (US) and python statsmodels v0.12.0 (Israel, UK).

### Country-level testing and case data

We reviewed publicly available data^15,21^ regarding testing guidelines in each region during the study period. We specifically sought information regarding the shift in testing criteria from core CLI symptoms (i.e. fever, respiratory symptoms) to a broader list of CLI symptoms. Open testing started on March 14th 2020 in the US while broader symptom-based testing occurred later in the UK (May 18th, 2020) and Israel (June 1st, 2020) ^15,19^. In addition, these dates coincided with inclusion of anosmia/ageusia, except for the US (April 5th, 2020).

## Results

### National COVID-19 surveillance platform participants

The study users and survey respondents compared to national demographics are shown (Table 1). Those participating in technology-based, health-related surveys tend more often to be female, younger, and healthier than the general population^22–24^, and this trend is borne out in these surveillance platforms. Survey-weighted US cohort data was more representative of the US (Supplementary table 2), but use of survey weights had little effect on results (Supplementary figure 2). The unweighted data analyses are presented throughout to be consistent across platforms.

**Table 1.** Baseline characteristics of national platform users and survey respondents in relation to national government demographics from cbs.gov.il, ONS.gov.uk, and census.gov (2019 estimates). See Supplementary table 2 for US data using survey weights. For US data were result to test was not always available, resulting data is indicated in parenthesis

|  | Israel |  | United Kingdom |  | United States |  |
| --- | --- | --- | --- | --- | --- | --- |
|  | Survey | Country | App | Country | Survey | Country |
| Number (adults only) | 15,550 | 6,015,116 | 3,360,949 | 44,105,500 | 6,626,897 | 255,200,373 |
| Age (adults only) | 60.2 (15.9) | 45.3 | 45.3 (15.6) | 47.7 | 48.5 | 47.8 |
| Male (%) | 50.9 | 49.6 | 38.5 | 49.5 | 33.3 | 49.5 |
| Tests | 48,571 | 1,774,736 | 341,500 | 9,415,384 | 238,316 (resulted 199,192)* | 62,092,416 |
| Positive tests | 84 | 70,379 | 7,030 | 302,301 | 28,355 | 4,495,014 |

### Testing capacity during the fall and rise of COVID-19 cases

During the study period (April through July 2020), SARS-CoV-2 testing capacity was being scaled up (Figure 1a). Meanwhile, government-reported COVID-19 cases declined after April 2020 (the “first wave” peak) due to a combination of interventions^25^. COVID-19 cases recredesced, first in Israel, and then in the US (Figure 1b). This “second wave” took place after the study period in the UK. National testing data are consistent with platform-specific testing data (Figure 1c). Additionally, UK platform-invited testing of mildly symptomatic (Supplementary figure 1) in early May 2020 was followed by nationally-mandated expansion.

**Figure 1.**
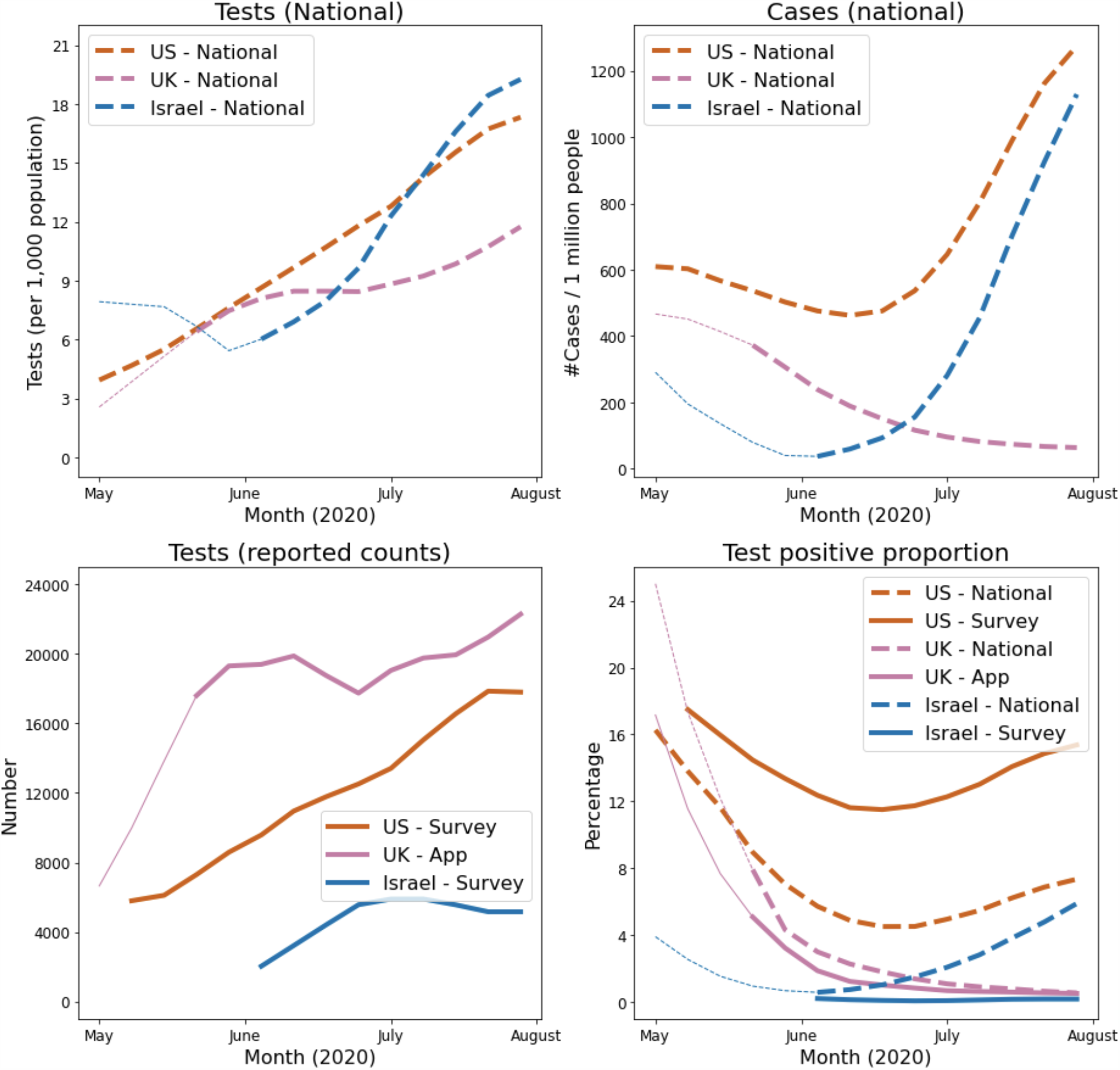
Comparison of the weekly (top left) tests per capita by country, (top right) cases per capita by country, (bottom left) test results by platform and (bottom right) test positive proportion by country and platform reported by platform during the study period in Israel (blue), UK (purple) and US (red). National data shown as dashed lines while surveillance platform data (bottom panels) shown as solid lines. Transition from thin to thick lines when testing policies were considered open.

The test positive proportion in the government and national surveillance platform trends (Figure 1d) were generally similar to each other, and to the case data. For the UK, the test positive proportion remained lower for longer due to rising testing and falling cases. US survey test proportion was higher but the trend is representative (see Supplementary figure 2 and 3 for weights, incident/prevalent and outlier sensitivity analysis). While there was a strong national-platform test positive proportion agreement in the US and UK (root mean square error of 0.06 and 0.03, respectively), there was a discrepancy in the rise in Israel in the first weeks after introduction of the testing report question, though testing counts were lower.

### Covid-like-symptoms and SARS-CoV-2 test positivity over time

Symptom performance as measured by the age- and sex-adjusted OR for the primary outcome of test positive versust test negative was not constant (Figure 2, top). However, several symptoms showed consistently strong associations with test positivity across platforms and over time. These include core CLI components (fever with respiratory symptoms) that were in the initial World Health Organization CLI definition, as well as the unusual symptom of anosmia/ageusia (Figure 2, left).

**Figure 2.**
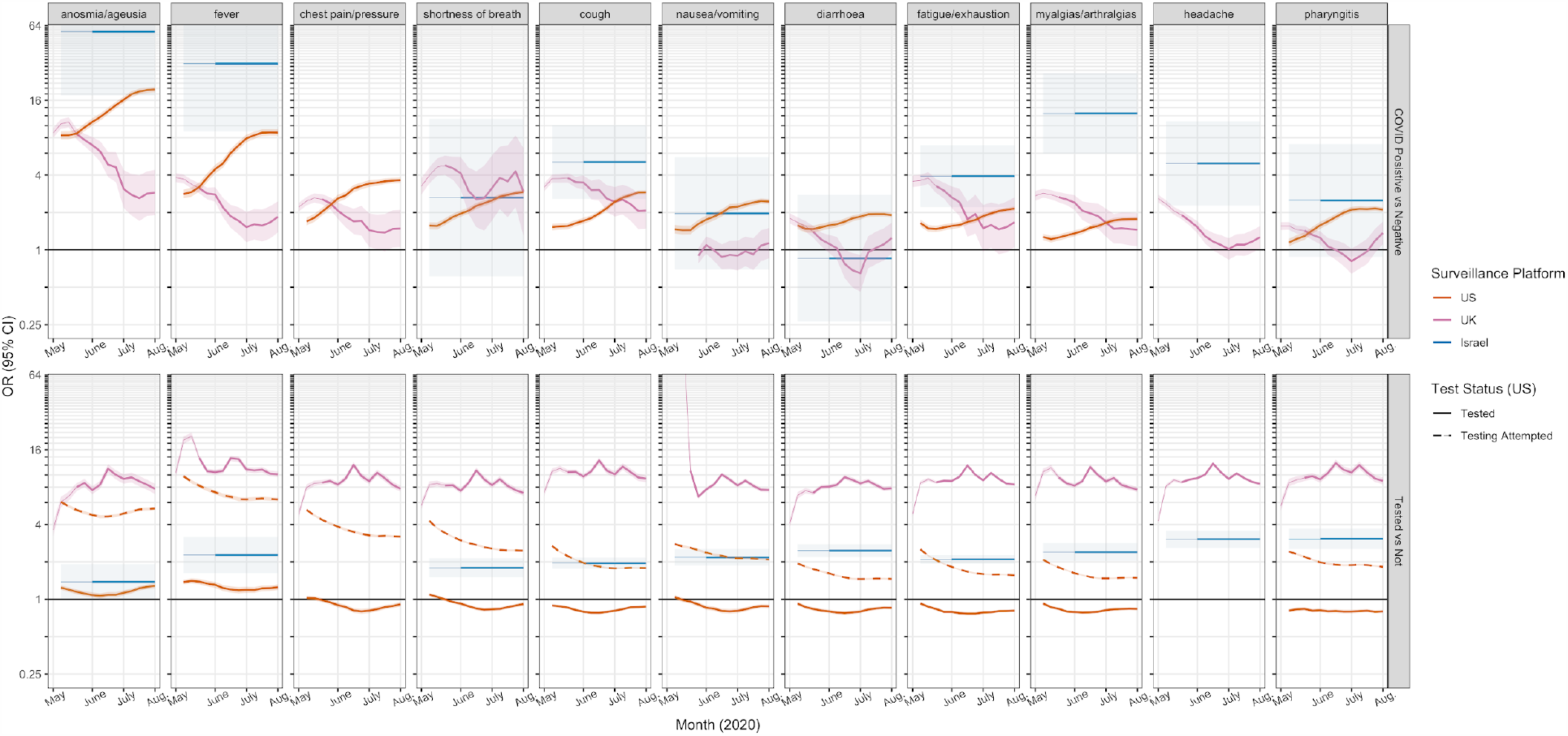
Comparison of odds ratios (OR) with 95% confidence intervals by platform for the outcome of test result positive versus negative (top facet), or tested versus not (bottom facet). Additionally, in the US platform, we show attempted testing versus not (dashed line), and tested versus not among those who attempted (solid line). Results from platforms for each country of Israel (blue), UK (purple) and US (red) are shown. Symptoms shared by at least two platforms in facet columns (see Supplementary material for sensitivity analyses, mapping and survey language). OR scale log-linear to enable comparisons across a wide range of effect estimates. Estimates calculated weekly using 5-week moving windows. Thin and thick lines indicate stricter versus broader symptoms for national testing practices (*NB*: prior to mid-June, the 5-week bins contain data from the stricter testing era).

Anosmia/ageusia was, at all times, across all platforms, the symptom with single highest OR for self-reported SARS-CoV-2 test positivity. The minimum OR (2.61 [95% confidence interval [1.71;3.98]) was during the lowest incidence following the 18th May 2020 inclusion of anosmia to UK testing criteria. Broader testing criteria and a rise in cases in the US (Figure 2) was coincident with a rising OR for many symptoms. and Israel (Supplementary Figure 5).

While CLI symptom signals were positive and relatively similar, gastrointestinal symptoms were equivocal in the UK and Israel. In the US, when restricting to oligosymptomatic surveys (≤ 5 symptoms self-reported, Supplementary figure 2), nausea and diarrhea, along with myalgias/arthralgias and pharyngitis, were no longer predictive of test-positivity. We hypothesize these findings may be due to clustering of symptoms or the phase of illness when testing completed.

Low incidence generally coincided with wider confidence intervals (see Figure 1). The US sampling scheme^20^ may have contributed to the more stable precision, though the timing of the tests relative to symptoms cannot be ascertained. To evaluate whether symptoms-to-test timing, illness duration or recall bias (e.g. US test and symptoms surveyed simultaneously) affects symptom signals, we conducted sensitivity analyses (Supplementary figure 3 and 4). While inclusion of post-testing symptoms generally resulted in higher effect estimates, especially anosmia/ageusia, chest pain/pressure, and fatigue/exhaustion, the temporal patterns of symptoms were similarly consistent.

### Covid-like-symptoms and SARS-CoV-2 testing access over time

The Covid-like symptoms of anosmia/ageusia, fever, cough and shortness of breath were the symptoms most strongly associated with obtaining SARS-CoV-2 testing (by self-report), regardless of test result (Figure 2, bottom).

Broader testing access and higher incidence (US) was concomitant with higher predictive power of symptoms. Symptoms became less predictive of test positivity and confidence intervals widened substantially in the UK with sustained low incidence.

Within the US platform, we assess differences in test seeking behavior from test access (among those who attempted to get testing). Early in the study period, anosmia/ageusia and core CLI drove self-reported test access, while test seeking generally paralleled the strength of the association with test positivity. UK test access was higher, in part because of platform-invited testing of mildly symptomatic users (Supplementary figure 1).

## Discussion

### Implications of key findings

Here we show convincing evidence that anosmia/ageusia is robustly associated with self-reported SARS-CoV-2 test positivity, regardless of the surveillance platform, country, testing guidelines, or pandemic phase. Overall, anosmia/ageusia was an order of magnitude more common among those reporting positive (US 43%, UK 44%, Israel 13.9%) compared to negative (US 5%, UK 3%, Israel 0.17%). This finding supports test access and self-isolation mandates with anosmia/ageusia^19,21,26^.

Core CLI components of fever and respiratory symptoms performed well. Importantly, symptom associations changed with testing expansion and case burden, while some were inconsistent predictors. These findings highlight key COVID-19 symptoms for multi-regional syndromic surveillance signals. Furthermore, there is a mandate to discriminate COVID-19 from seasonal respiratory pathogens like influenza^27^, though there are few data sets^1,2^ from which to define discriminating symptoms *a priori*. A prospective, iterative, surveillance-data based approach, leveraging national platforms such as these, is likely to play an important role.

These findings show the power of citizen science in the COVID-19 response. Though privacy limits validation of anonymous self-reports against health records, the near-real-time survey-based outcomes closely mirror national trends, and are therefore useful for “nowcasting” and forecasting ^28,29^. They also demonstrate the utility of early broad-based testing in conjunction with population-wide surveillance. Indeed, testing is a cornerstone of pandemic response, and has presented substantial challenges globally ^10,16^. We show the impact of differences in test seeking or access (e.g. core CLI symptoms) and low prevalence (e.g. larger confidence intervals) on univariate disease prediction. Future directions that build on these findings include the incorporation of cross-platform, spatio-temporal and and multivariate effect estimates in the development of global prediction public health tools.

A strength of digital participatory surveillance is that data collection can be initiated rapidly and tailored to public health needs (e.g. longitudinal disease trajectory, consistent or representative population sampling) over space and time. However, to conduct this inter-platform international comparison of symptom-based COVID-19 prediction, we had to map survey questions (e.g. subjective fever versus temperature threshold) and account for study design variation (e.g. US cross-sectional simultaneously queried symptoms over prior 24 hours and any test result, UK included symptom logged 14 days prior to test report). Sensitivity analyses show our findings are robust to relaxing assumptions such as illness duration, symptom-to-test window, symptom report pattern, platform-suggested testing and the use of survey weights. The possibility to be tested multiple times over the course of the disease was beyond the scope of this study, and not feasible with one-time surveys. Symptoms that appear at a later disease stage (e.g. shortness of breath) had very wide confidence intervals, while symptoms that appear in conjunction with others (e.g. myalgias/arthralgias) were not predictive when filtering on few total symptoms.

### Limitations

These findings must be interpreted with the caveat that, by its nature, real-time participatory syndromic surveillance inherently has potential biases related to (a) generalizability (whether participants representative of the source population, or have covariates for critical effect modifiers), and (b) measurement bias (survey question misunderstanding, differential missingness or error in self-reporting due to incentive to log healthy when being monitored or misuse one-time surveys), as examples. We compared each platform to national demographics and outcomes, as well as survey-weighted outcomes in the US. For both the UK and US platforms, respondents were younger and more often female, similar to published online survey participation demographics and echoes research showing possible biases related to use of mobile health devices solutions in the context of symptom reporting in the COVID19 era ^22–24^. To address measurement bias, we compared symptom patterns and symptom windows across platforms; while these affected the magnitude of effect estimates, the overall trends and the strength of anosmia/ageusia and core CLI symptom in the prediction of COVID-19 held.

### Strengths

Despite these limitations, the strength of this study lies in the combination of data from very different digital platforms varied in terms of their participants’ location (Israel, UK, USA) and their observation over time (April to July 2020). All three datasets combined are very large in size (over 10 million respondents) with high numbers of tests (over half a million), and the capacity to provide automated, aggregate outcomes in near-real time. In addition, the variety in access to test and healthcare policies as well as the differences in epidemic timeline across countries make these findings more generalisable.

## Conclusion

To our knowledge, this is the first cross-country, cross-platform temporal comparison of predictive symptoms of COVID-19. We demonstrate the generalisability of the unique symptom of anosmia/ageusia. In addition, the strength of CLI symptoms to predict test positivity will vary with test access, incidence and phase of illness. This confirmed the strength of fever and respiratory symptoms as good CLI signals, and importantly strongly supports early, broad-based, community-wide testing in a pandemic.

## Data Availability

Israel Platform
Tables of de-identified, aggregated data are available at https://github.com/hrossman/Covid19-Survey.
UK Platform
Data used in this study is available to bona fide researchers through UK Health Data Research using the following link
https://web.www.healthdatagateway.org/dataset/fddcb382-3051-4394-8436-b92295f14259
US Platform:
Requests for access to the US Carnegie Mellon University/ Facebook COVID-19 Symptom Survey available via https://dataforgood.fb.com/docs/covid-19-symptom-survey-request-for-data-access/.

## Acknowledgments

CMA: National Institutes of Health K23 DK120899, Boston Children’s Hospital Office of Faculty Development Career Development Award. CHS: Alzheimer’s Society Junior Fellowship - AS-JF-17-011, ADJ: NIH K01 DK 110267, EM: ‘Skills Development Scheme’ of the Medical Research Council UK. Support for the Covid Symptom Study (UK data) was provided by the NIHR-funded Biomedical Research Centre based at GSTT NHS Foundation Trust. This work was supported by the UK Research and Innovation London Medical Imaging & Artificial Intelligence Centre for Value Based Healthcare.

## Data Availability

### Israel Platform

Tables of de-identified, aggregated data are available at https://github.com/hrossman/Covid19-Survey.

### UK Platform

Data used in this study is available to bona fide researchers through UK Health Data Research using the following link
https://web.www.healthdatagateway.org/dataset/fddcb382-3051-4394-8436-b92295f14259

### US Platform

Requests for access to the US Carnegie Mellon University/ Facebook COVID-19 Symptom Survey available via https://dataforgood.fb.com/docs/covid-19-symptom-survey-request-for-data-access/.

## Supplementary material

**Supplementary figure 1:**
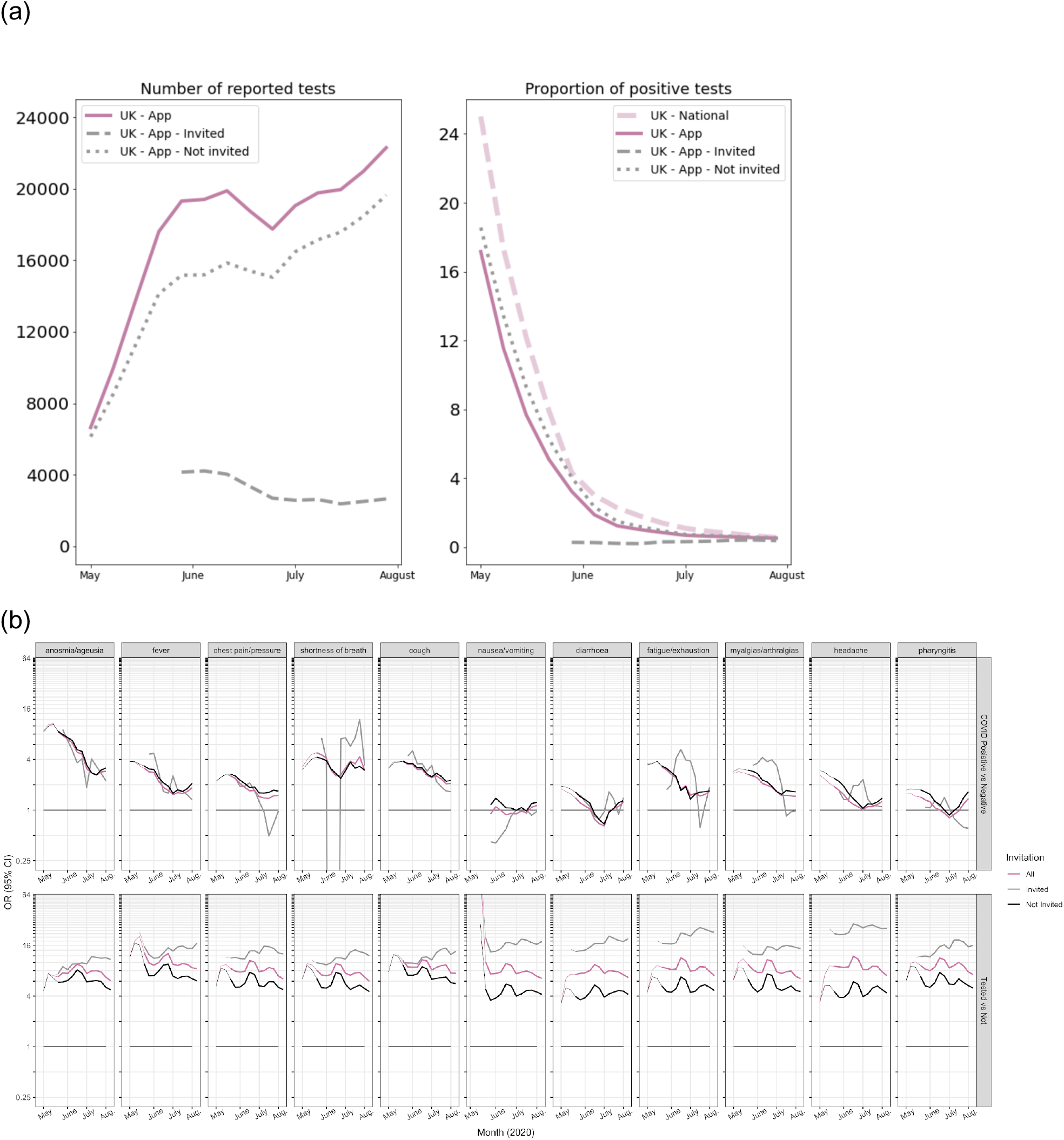
UK invited testing sensitivity analysis UK platform primary analysis, invited-test respondents and not invited-test respondents. For those with repeated tests, negative test results following a positive test were censored. In the UK study, beginning 29 April 2020, oligosymptomatic individuals were invited to participate in platform-sponsored SARS-CoV-2 PCR testing, regardless of whether they met testing guidelines in the region. Invited data are plotted starting from the first 5-week bin, while the full data are plotted from study inception (which includes increasing numbers of invited tests).

**Supplementary figure 2:**
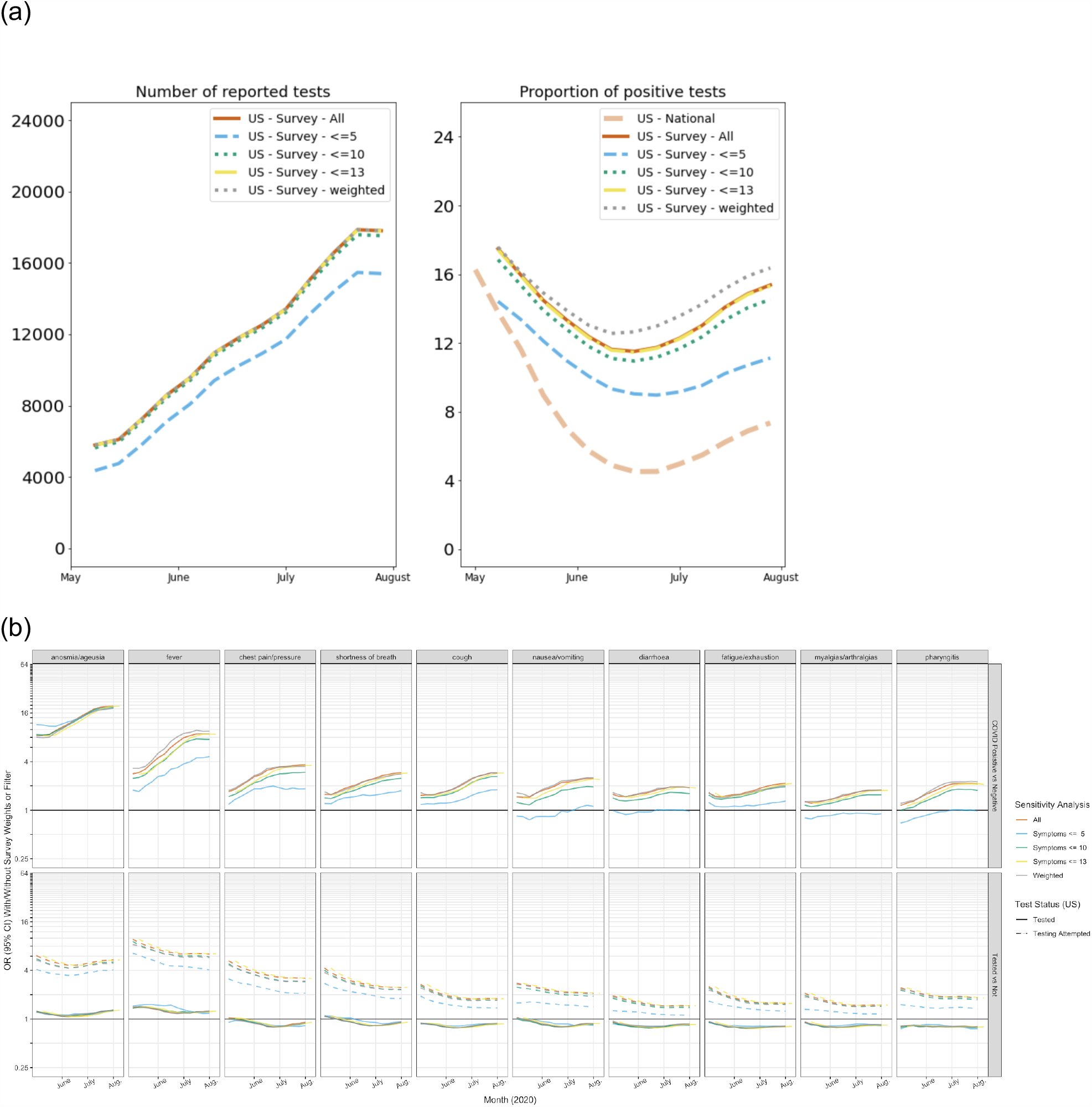
US weights and filtering sensitivity analysis US platform (Facebook Symptom Survey) OR (95% confidence interval) effect estimates from the main analysis, alongside sensitivity analyses with strict filtering (including surveys with no more than 5, 10 and 13 of 14 symptoms reported during the prior 24 hours), or with survey weights (Facebook Active User Base “raked” to the US census population)^30^.

**Supplementary figure 3:**
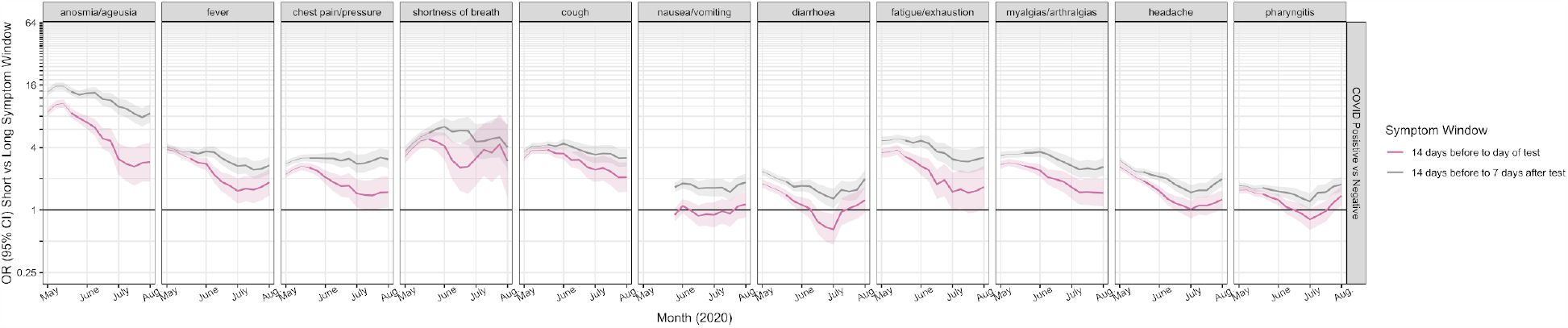
UK symptom-to-test window sensitivity analysis UK (Zoe Covid Study) Short versus long window for symptoms, with long window including symptom up to 7 days following the test report. Short window as shown in the main figure includes 14 days preceding the test report.

**Supplementary figure 4:**
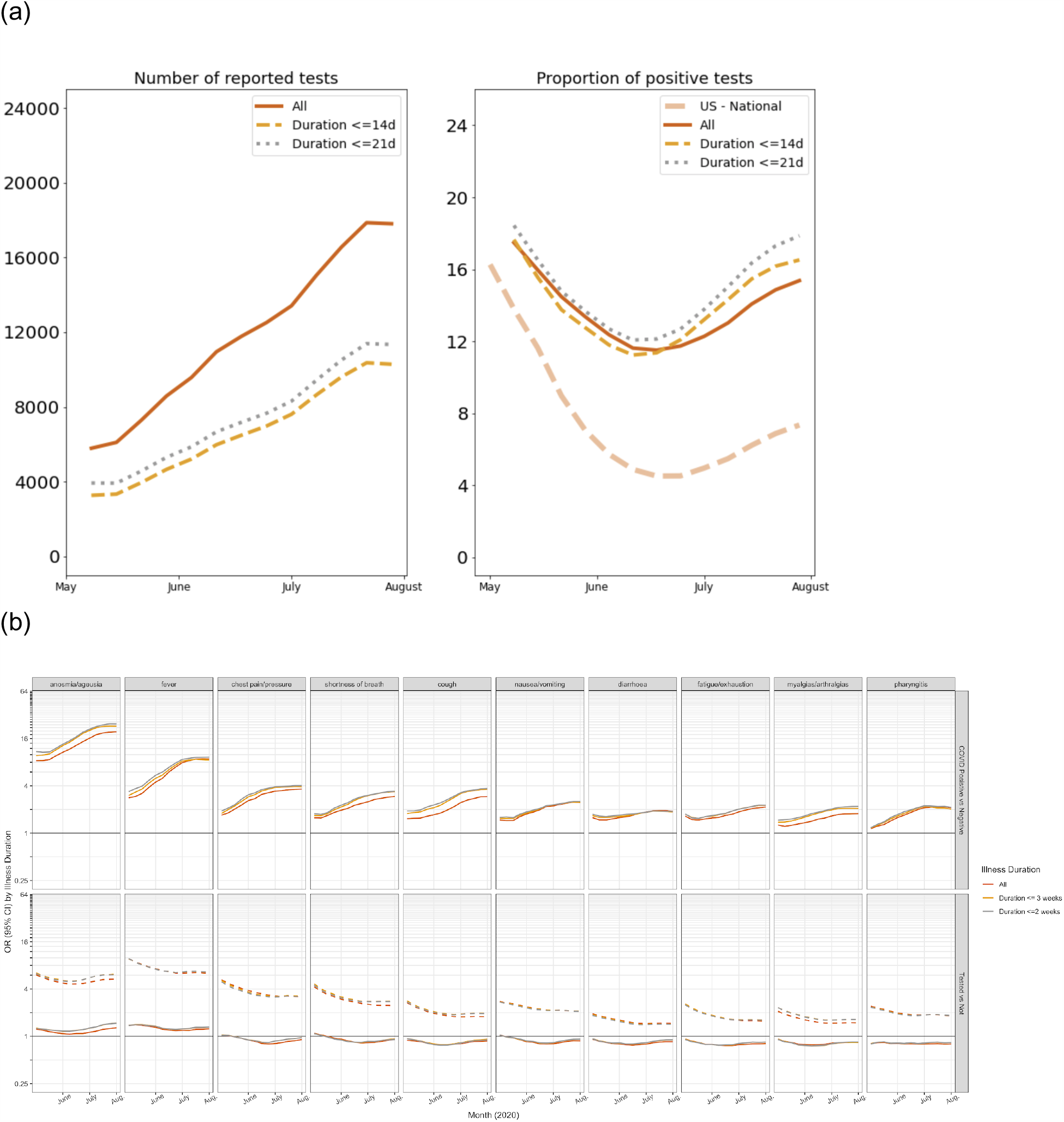
US illness duration sensitivity analysis US (Facebook Symptom Survey) Self-reported duration of illness of any length compared to 14 and 21 days.

**Supplementary Figure 5:**
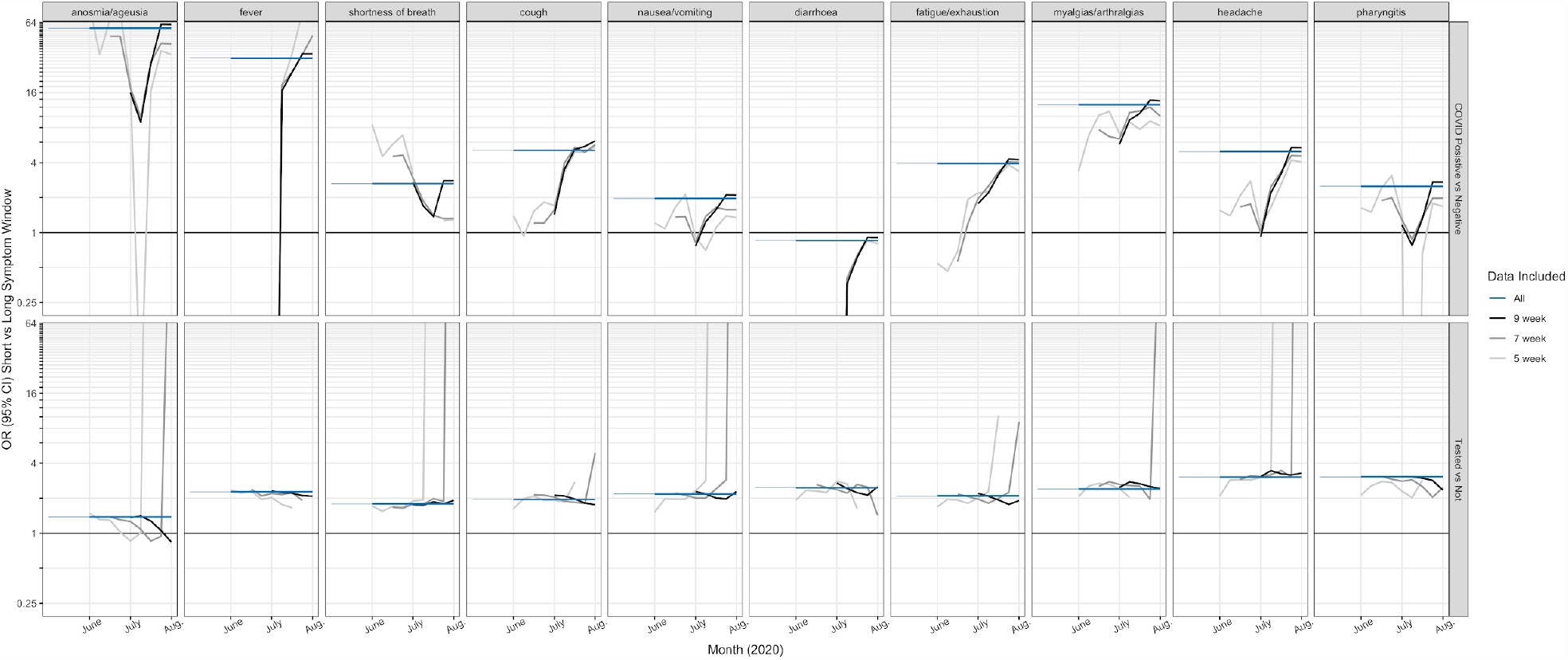
Increasing windows for Israel data aggregation Israel Corona data aggregated over 5-, 7- and 9-week windows to show trends with expansion of testing (e.g. cough effect estimate increased). For the primary analyses, aggregate data over the entire study period are shown due to smaller sample size.

**Supplement table 1:**
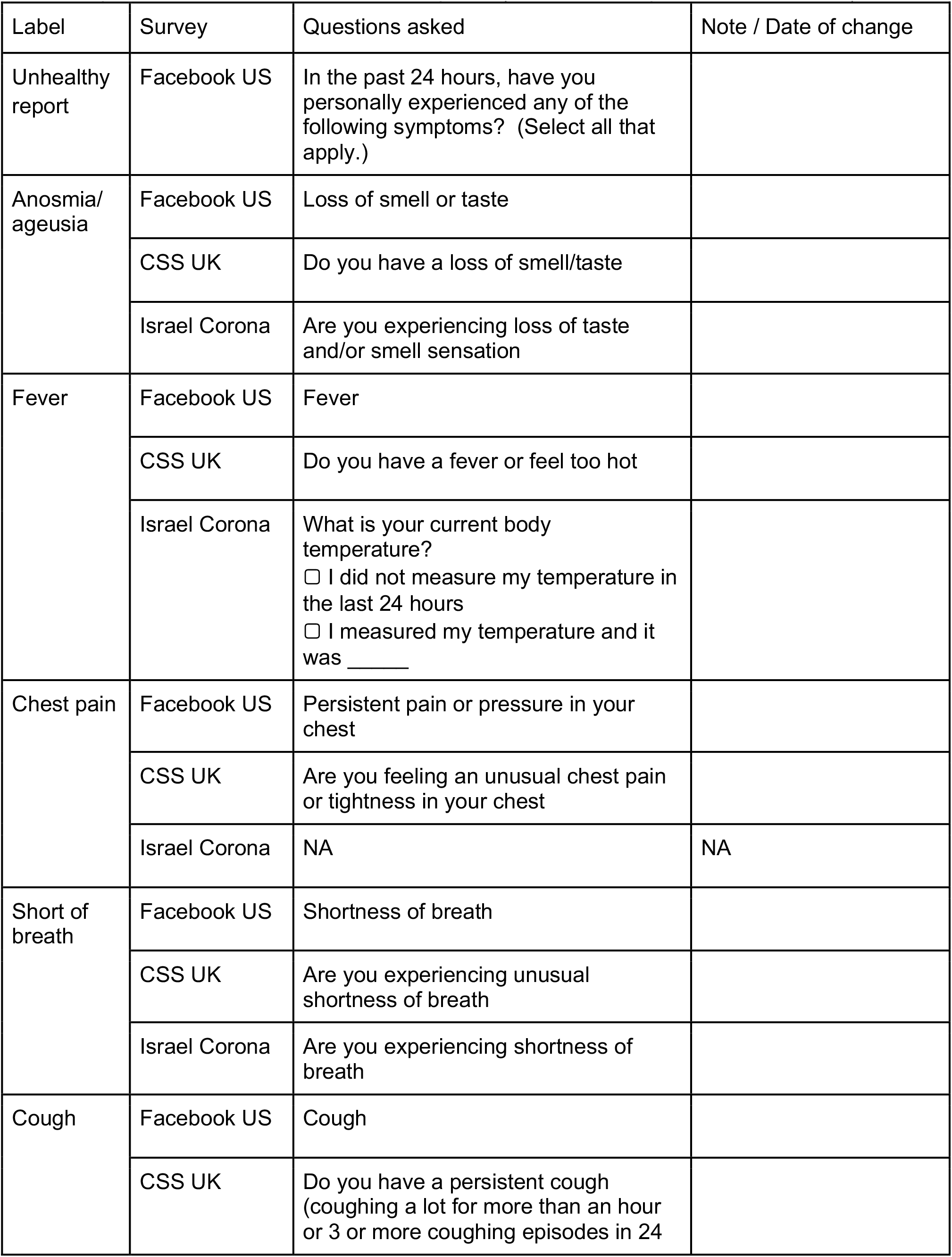
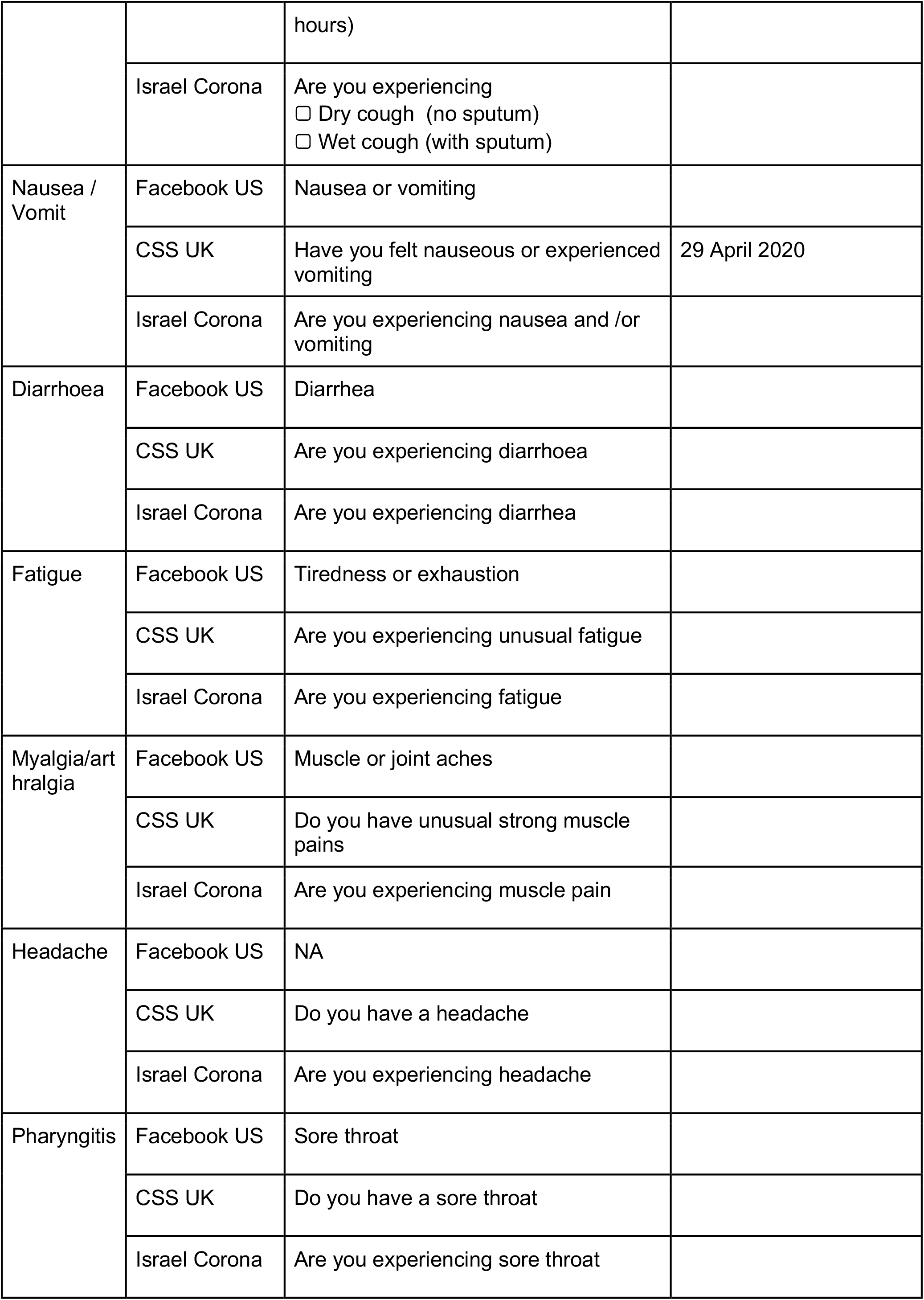
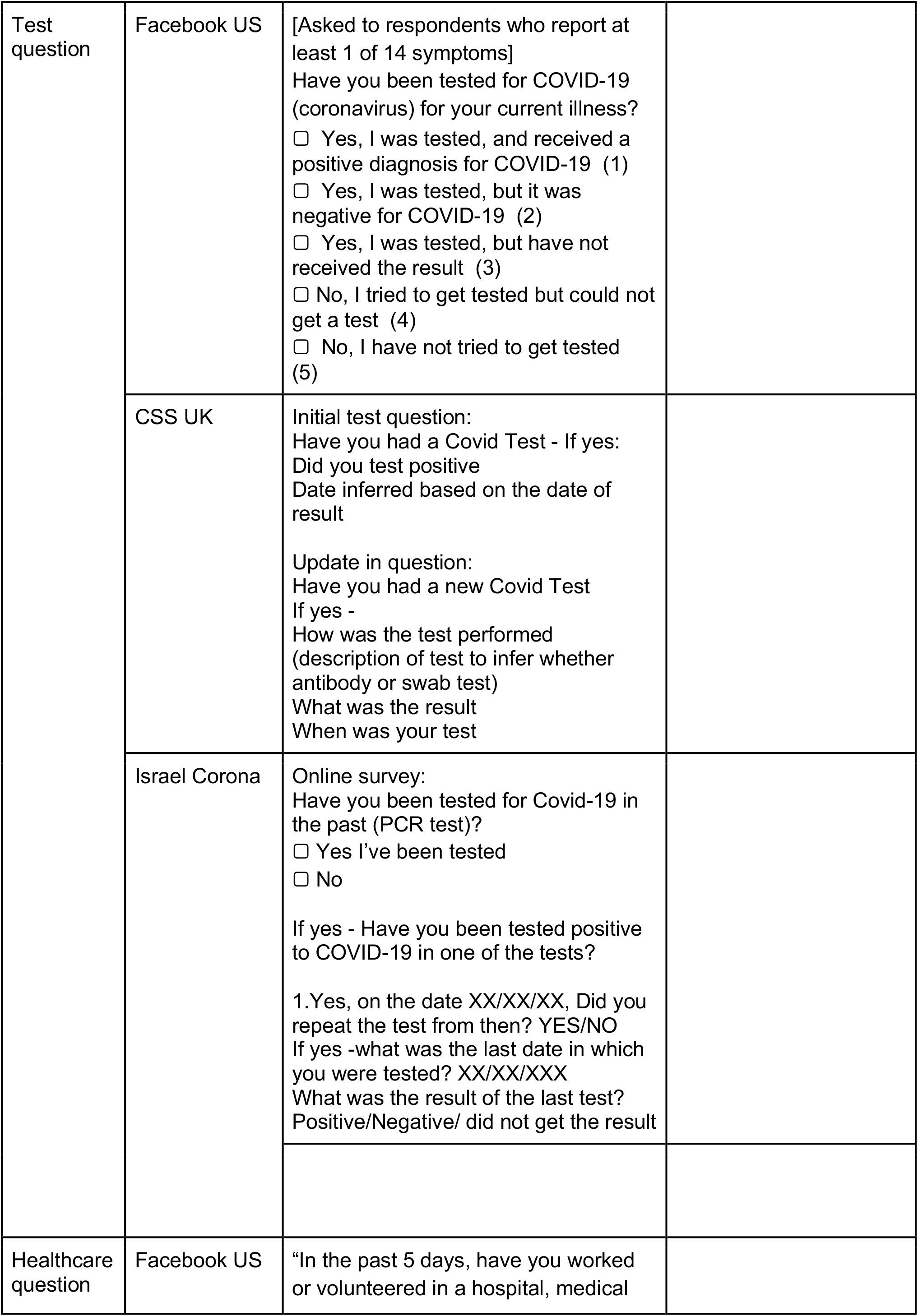
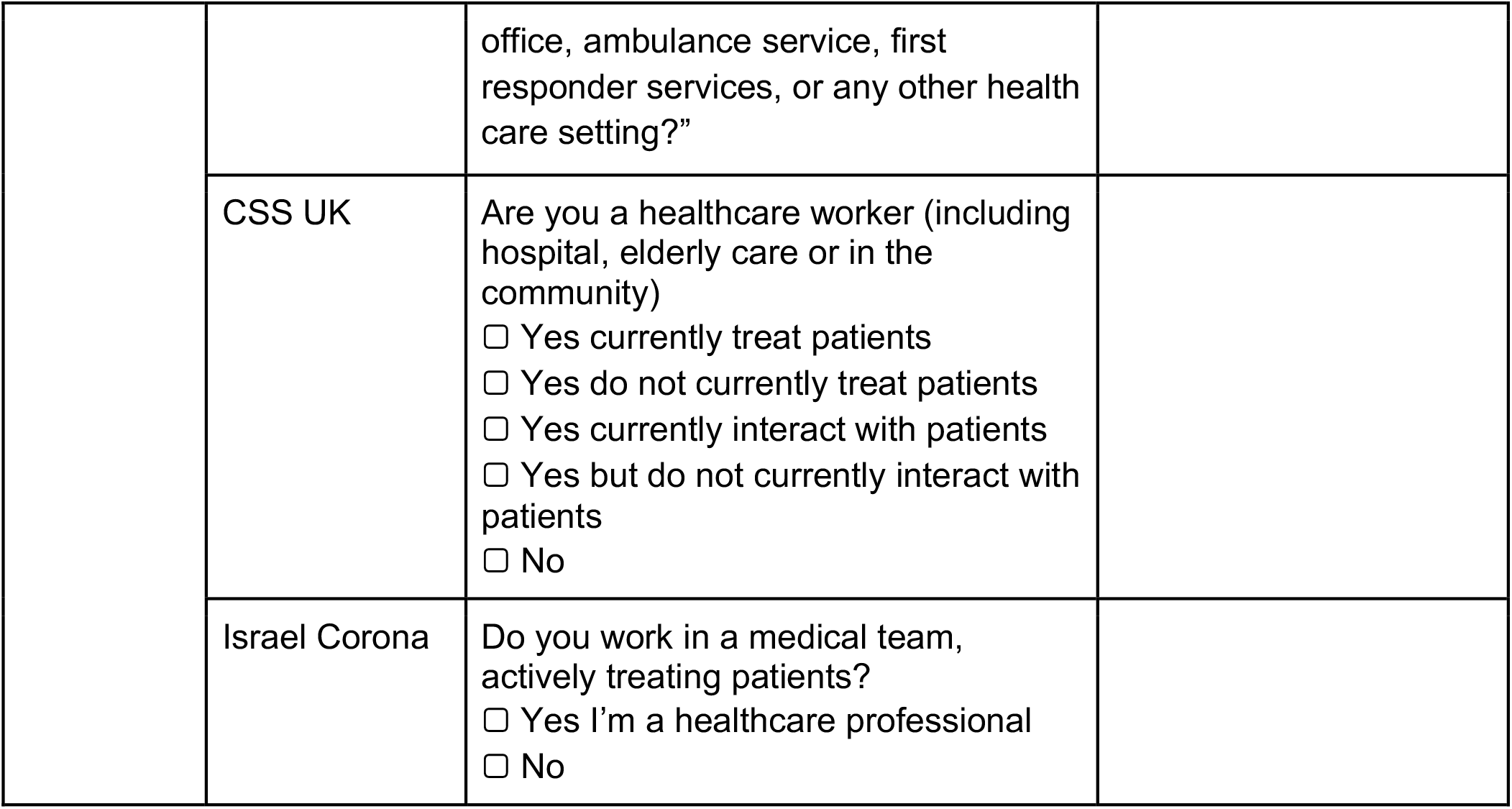
Mapping of survey questions across platforms Mapping of questions and symptoms across datasets. Eleven symptoms were shared across at least two of three platforms, after having excluded symptoms that were shared but had limited responses, and thus could not be compared (i.e. abdominal pain, rash, confusion).

**Supplementary table 2:** Weight demographic characteristics for US platform Baseline demographic characteristics of non-healthcare worker US platform survey respondents age ≥ 18 years reporting male or female sex, in relation to the US population, with and without application of survey weights.

|  | United States |  |  |
| --- | --- | --- | --- |
|  | Raw Survey | Survey-Weighted | Country |
| Age (adults only) | 48.5 | 48.7 | 47.8 |
| Sex | 33.3 | 48.1 | 49.5 |

## References

1 Smolinski MS, Crawley AW, Baltrusaitis K, et al. Flu Near You: Crowdsourced Symptom Reporting Spanning 2 Influenza Seasons. Am J Public Health 2015; 105: 2124–30.

2 Carlson SJ, Dalton CB, Durrheim DN, Fejsa J. Online Flutracking survey of influenza-like illness during pandemic (H1N1) 2009, Australia. Emerg Infect Dis 2010; 16: 1960–2.

3 Barkay N, Cobb C, Eilat R, et al. Weights and Methodology Brief for the COVID-19 Symptom Survey by University of Maryland and Carnegie Mellon University, in Partnership with Facebook. arXiv [cs.SI]. 2020; published online Sept 25. http://arxiv.org/abs/2009.14675.

4 Drew DA, Nguyen LH, Steves CJ, et al. Rapid implementation of mobile technology for real-time epidemiology of COVID-19. Science 2020; published online May 5. DOI:10.1126/science.abc0473.

5 Rossman H, Keshet A, Shilo S, et al. A framework for identifying regional outbreak and spread of COVID-19 from one-minute population-wide surveys. Nat Med 2020; 26: 634– 8.

6 Allen WE, Altae-Tran H, Briggs J, et al. Population-scale longitudinal mapping of COVID-19 symptoms, behaviour and testing. Nat Hum Behav 2020; 4: 972–82.

7 Budd J, Miller BS, Manning EM, et al. Digital technologies in the public-health response to COVID-19. Nat Med 2020; 26: 1183–92.

8 Jansen-Kosterink SM, Hurmuz M, den Ouden M, van Velsen L. Predictors to use mobile apps for monitoring COVID-19 symptoms and contact tracing: A survey among Dutch citizens. medRxiv 2020. https://www.medrxiv.org/content/10.1101/2020.06.02.20113423v1.abstract.

9 CovidNearYou. https://covidnearyou.org/ (accessed Dec 11, 2020).

10 Lipsitch M, Swerdlow DL, Finelli L. Defining the Epidemiology of Covid-19 - Studies Needed. N Engl J Med 2020; 382: 1194–6.

11 The COVID Tracking Project. https://covidtracking.com/ (accessed Dec 11, 2020).

12 Griffith GJ, Morris TT, Tudball MJ, et al. Collider bias undermines our understanding of COVID-19 disease risk and severity. Nat Commun 2020; 11: 5749.

13 Lipsitch M, Donnelly CA, Fraser C, et al. Potential Biases in Estimating Absolute and Relative Case-Fatality Risks during Outbreaks. PLoS Negl Trop Dis 2015; 9: e0003846.

14 Menni C, Valdes AM, Freidin MB, et al. Real-time tracking of self-reported symptoms to predict potential COVID-19. Nat Med 2020; published online May 11. DOI:10.1038/s41591-020-0916-2.

15 Oxford COVID-19 Government Response Tracker. https://covidtracker.bsg.ox.ac.uk/ (accessed Dec 11, 2020).

16 Rader B, Astley CM, Sy KTL, et al. Geographic access to United States SARS-CoV-2 testing sites highlights healthcare disparities and may bias transmission estimates. J Travel Med 2020; 27. DOI:10.1093/jtm/taaa076.

17 Organization WH, Others. Global surveillance for COVID-19 caused by human infection with COVID-19 virus: interim guidance, 20 March 2020. World Health Organization, 2020 https://apps.who.int/iris/bitstream/handle/10665/331506/WHO-2019-nCoV-SurveillanceGuidance-2020.6-eng.pdf.

18 Gandhi RT, Lynch JB, Del Rio C. Mild or Moderate Covid-19. N Engl J Med 2020; 383: 1757–66.

19 Council of State and Territorial Epidemiologists. Standardized surveillance case definition and national notification for 2019 novel coronavirus disease (COVID-19). 2020.

20 Delphi and Facebook COVID Symptom Survey. https://cmu-delphi.github.io/delphi-epidata/symptom-survey/ (accessed Nov 17, 2020).

21 State of Israel Ministry of Health COVID-19 information. https://govextra.gov.il/ministry-of-health/corona/corona-virus-en/ (accessed Nov 17, 2020).

22 Baltrusaitis K, Santillana M, Crawley AW, Chunara R, Smolinski M, Brownstein JS. Determinants of Participants’ Follow-Up and Characterization of Representativeness in Flu Near You, A Participatory Disease Surveillance System. JMIR Public Health and Surveillance. 2017; 3: e18.

23 Korkeila K, Suominen S, Ahvenainen J, et al. Non-response and related factors in a nation-wide health survey. Eur J Epidemiol 2001; 17: 991–9.

24 Shaver LG, Khawer A, Yi Y, et al. Using Facebook Advertising to Recruit Representative Samples: Feasibility Assessment of a Cross-Sectional Survey. J Med Internet Res 2019; 21: e14021.

25 Flaxman S, Mishra S, Gandy A, et al. Estimating the effects of non-pharmaceutical interventions on COVID-19 in Europe. Nature 2020; 584: 257–61.

26 COVID-19: investigation and initial clinical management of possible cases. UK Government Guidance. https://www.gov.uk/government/publications/wuhan-novel-coronavirus-initial-investigation-of-possible-cases/investigation-and-initial-clinical-management-of-possible-cases-of-wuhan-novel-coronavirus-wn-cov-infection#criteria.

27 Rubin R. What Happens When COVID-19 Collides With Flu Season? JAMA 2020; 324: 923–5.

28 COVID-19 Symptom Surveys through Facebook. https://delphi.cmu.edu/blog/2020/08/26/covid-19-symptom-surveys-through-facebook/..

29 Varsavsky T, Graham MS, Canas LS, et al. Detecting COVID-19 infection hotspots in England using large-scale self-reported data from a mobile application: a prospective, observational study. The Lancet Public Health 2020; published online Dec 3. DOI:10.1016/S2468-2667(20)30269-3.

30 Delphi and Facebook COVID Symptom Survey Weights. https://cmu-delphi.github.io/delphi-epidata/symptom-survey/symptom-survey-weights.pdf (accessed Nov 18, 2020).

